# Beyond Binary MRD: Quantitative ctDNA Interpretation After Curative-Intent Surgery for Colorectal Cancer

**DOI:** 10.64898/2026.03.09.26347910

**Authors:** Jungmin Kim, Sunghyeok Ye, Jung-Myun Kwak, Dahee Choi, Suhyeon Kim, Ho Jin Jeong, Eunju Hong, Jae Woo Lee, Sujin Kim, Yoon-Ho Won, Sung Sun Koo, In Seon Lee, Taegun Park, Ju Bin Yoon, Harim Oh, Yoo Jin Lee, Sang-Jung Ahn, Jin-Soo Kim, Hong-Kyu Kim, Hyun-Woong Cho, Sanghoo Lee, Jeonghoon Hong, Pedram Razavi, Jin Kim, Junseok W Hur

## Abstract

**Background:** Circulating tumor DNA (ctDNA) detection after curative-intent surgery is being used to identify minimal residual disease (MRD) in colorectal cancer (CRC). However, MRD classification is dependent on analytical sensitivity, and the impact of detection threshold on observed post-operative positivity remains incompletely characterized. We evaluated MRD positivity in stage I–III CRC using a CRISPR-based plasma sequencing assay, MUTE-Seq.

**Methods:** Patients were prospectively enrolled and analyzed using customized tumor-informed panels applied to baseline and post-operative plasma samples collected at 4-week and 3-month. We report preliminary results from 39 plasma samples obtained from the first 14 patients. MRD positivity was assessed across multiple hypothetical detection thresholds (1–100 ppm).

**Results:** All 14 patients (100%) had detectable mutations at baseline. Mutation-positive call number significantly decreased after surgery (baseline vs 4-week, p = 0.006; baseline vs 3-month, p = 0.004), and ctDNA concentration likewise declined (baseline vs 4-week, p = 0.002; baseline vs 3-month, p = 0.003). Among stage II–III patients, MRD positivity at 4-week was 20% at a 100-ppm threshold but increased to 70% at 10 ppm and 100% at 1 ppm. At 3-month, MRD positivity was 11% at a 100-ppm threshold and 78% at 1 ppm. At both time points, approximately 80% of MRD-positive stage II–III patients harbored ctDNA levels below 100 ppm, and half of these cases were below 15 ppm. Two patients (one stage I and one stage II) developed recurrence; both were MRD-positive at 4-week and demonstrated increasing mutation-positive calls at 3-month, with a median radiologic lead time of 4 months.

**Conclusions:** Post-operative MRD classification in CRC is strongly influenced by analytical sensitivity. A substantial proportion of residual disease signals reside below the conventional ctDNA detection threshold of 100 ppm, supporting the clinical relevance of ultrasensitive ctDNA detection.

## Introduction

Accounting for more than 150,000 new cases and 50,000 deaths annually in the United States, Colorectal cancer (CRC) is the second leading cause of cancer-related death worldwide (*1-3*). Despite improvements in screening and advances in treatment, disease recurrence remains frequent. Up to 35% of patients with localized CRC eventually relapse, and the 5-year mortality rate remains around 40% (*4, 5*).

Surgical resection is the primary treatment for patients with stages I to III CRC, with adjuvant chemotherapy (ACT) recommended for selected patients (*6*). However, current criteria for ACT selection have room for improvement. Most patients with stage II disease do not receive ACT, although approximately 10% to 15% have clinically undetectable tumor remnants after surgery and are at risk for recurrence (*7*). Conversely, ACT is routinely administered to patients with stage III CRC, even though more than 50% are cured by surgery alone, while around 30% still relapse despite treatment (*8-11*). These limitations indicate the need for more precise tools to identify patients at high risk of recurrence and to better determine which individuals are most likely to benefit from adjuvant therapy, thereby informing clinical decision-making.

Plasma-based analysis of circulating tumor DNA (ctDNA) has emerged as a promising approach for detecting minimal residual disease (MRD) and monitoring disease longitudinally (*12*). Advances in molecular technologies now allow detection of very low variant allele fractions (VAFs), enabling identification of MRD after curative-intent treatment. Released into the bloodstream by tumor cells, ctDNA has a short half-life of around 1 hour, allowing near real-time assessment of disease status (*13*). In CRC, post-operative ctDNA positivity has been reported in around 10% to 50% of patients, depending on disease stage, and ctDNA detection can precede radiologic recurrence by a median of around 8 to 11 months (*14-16*).

However, the utility of ctDNA for detecting MRD depends strongly on analytical sensitivity. Assays with limited sensitivity may fail to detect residual tumors and lead to missed opportunities for early treatment (*17, 18*). In one study of resected CRC, a ctDNA assay identified recurrence in 53.3% of cases, which was comparable to detection by imaging and carcinoembryonic antigen (CEA), with a detection rate of 60.0%. Based on these findings, the authors concluded that the ctDNA assay did not provide a clear advantage over standard surveillance methods (*19*). In the MRD monitoring setting, it has been suggested that a limit of detection (LOD) of 0.01% VAF or lower is required to reliably identify residual disease and early recurrence (*20*). These observations indicate that high analytical sensitivity is essential for ctDNA-based approaches to inform clinical decision-making in colorectal cancer.

This study assessed the performance of customized panels using *Mutation Tagging by CRISPR-based Ultra-precise Targeted Elimination in Sequencing* (MUTE-Seq), a tumor-informed ultrasensitive ctDNA assay that enhances detection of low-level variants through selective depletion of wild-type DNA in the MRD setting (21). The assay was evaluated as a plasma-based approach for identifying trace ctDNA in patients with stage I–III CRC undergoing curative-intent surgery. Because post-operative MRD positivity depends strongly on assay sensitivity, we further examined how varying detection thresholds influence MRD positivity rates in stage II–III disease. Baseline and post-operative plasma samples from 14 patients were analyzed at the first (4-week) and second (3-month) follow-up visits, hereafter referred to as *Visit 1* and *Visit 2*, respectively. MRD positivity was assessed across multiple hypothetical detection thresholds. Results will assist comparison across assays with different analytical sensitivities and provide a more refined estimate of the proportion of patients with residual disease who may remain at risk for recurrence.

## Results

### Patient Characteristics

This study included 14 patients with stage I–III CRC undergoing curative-intent surgical resection. Median age at enrollment was 61.0 years (interquartile range [IQR], 51.0–73.0), and 9 patients (64.3%) were female. Four patients (28.6%) had stage I disease, 7 (50.0%) had stage II, and 3 (21.4%) had stage III disease. Most tumors were moderately differentiated adenocarcinomas (n = 13, 92.9%), with one case of mucinous adenocarcinoma (n = 1, 7.1%). Risk stratification by European Society for Medical Oncology (ESMO) criteria identified six of the seven stage II patients as low risk and one as high risk. Among the three stage III patients, one was classified as low risk and two as high risk. No patient had fewer than 12 lymph nodes assessed at the time of surgical resection. Patient demographics and disease characteristics are summarized in Table 1. One patient with stage I disease withdrew prior to *Visit 1*, and one patient with stage III disease withdrew prior to *Visit 2*. Accordingly, 13 patients were evaluable at *Visit 1* and 12 patients were included in the *Visit 2* analysis. Median follow-up period was 7.6 months.

**Table 1.** Patient Demographics and Disease Characteristics (N = 14)

| Characteristic |  | Value |
| --- | --- | --- |
| Age, years |  | 61.0 [51.0-73.0] |
| Sex |  |  |
| Female |  | 9 (64.3%) |
| Male |  | 5 (35.7%) |
| Tumor Location |  |  |
| Colon, unspecified |  | 2 (14.3%) |
| Sigmoid colon |  | 3 (21.4%) |
| Rectosigmoid |  | 1 (7.1%) |
| Rectum |  | 8 (57.1%) |
| Pathological Stage |  |  |
| I |  | 4 (28.6%) |
| II |  | 7 (50.0%) |
| ESMO Clinical Risk | Low | 6 (42.9%) |
|  | High | 1 (7.1%) |
| III |  | 3 (21.4%) |
| ESMO Clinical Risk | Low | 1 (7.1%) |
|  | High | 2 (14.3%) |
| Histology |  |  |
| Adenocarcinoma, moderately differentiated |  | 13 (92.9%) |
| Mucinous adenocarcinoma |  | 1 (7.1%) |
| BRAF status |  |  |
| N/A |  | 3 (21.4%) |
| Wild type |  | 11 (78.6%) |
| RAS status |  |  |
| N/A |  | 3 (21.4%) |
| KRAS wild type, NRAS wild type |  | 5 (35.7%) |
| KRAS wild type, NRAS mutated |  | 2 (14.3%) |
| KRAS mutated, NRAS wild type |  | 4 (28.6%) |

Values are presented as median [interquartile range] or n (%) *ESMO*, European Society for Medical Oncology; *N/A*, not applicable

### MRD Analysis Results and Clinical Outcomes

At baseline, all stage I–III patients had at least one detectable cancer-related mutation in plasma cfDNA. Across all stages, RAS/BRAF mutation status was not significantly associated with cfDNA mutation-positive call number (p = 0.747) or ctDNA levels (p = 0.592). Following surgical resection, the number of mutation-positive calls decreased across post-operative time points, as summarized in Table 2.

**Table 2.**
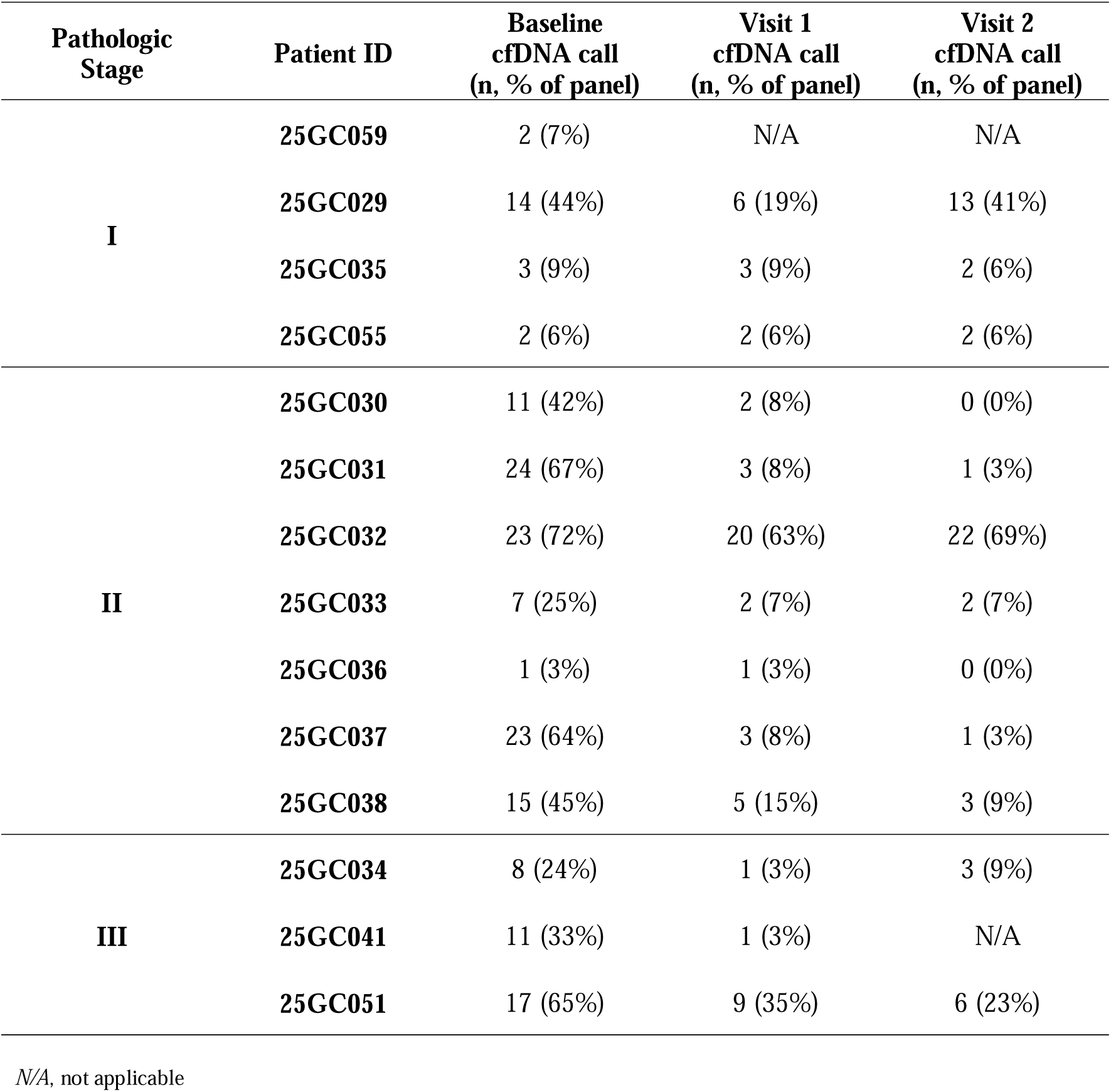
cfDNA Call at Baseline and Postoperative Follow-up.

The number of mutation-positive calls decreased significantly after surgery, from a median of 11 at baseline to 3 at *Visit 1* (p = 0.006) and to 2 at *Visit 2* (p = 0.004). The proportion of positive calls within the panel declined from 37.5% at baseline to 8% at *Visit 1* (p = 0.006). Likewise, ctDNA particle fraction (CPF; parts per million [ppm]) showed a marked reduction, decreasing from a median of 265.0 ppm (IQR, 62.90–595.00) at baseline to 20.8 ppm (IQR, 13.70–46.90) at *Visit 1* (p = 0.002) and 15.6 ppm (IQR, 7.63–37.00) at *Visit 2* (p = 0.003).

Between *Visit 1* and *2*, all ctDNA metrics continued to decline numerically, although these differences were not statistically significant (mutation positive call number, p = 0.531; panel-adjusted call proportion, p = 0.574; CPF, p = 0.108). Longitudinal changes in mutation-positive call number and CPF are illustrated in Figure 1.

**Figure 1.**
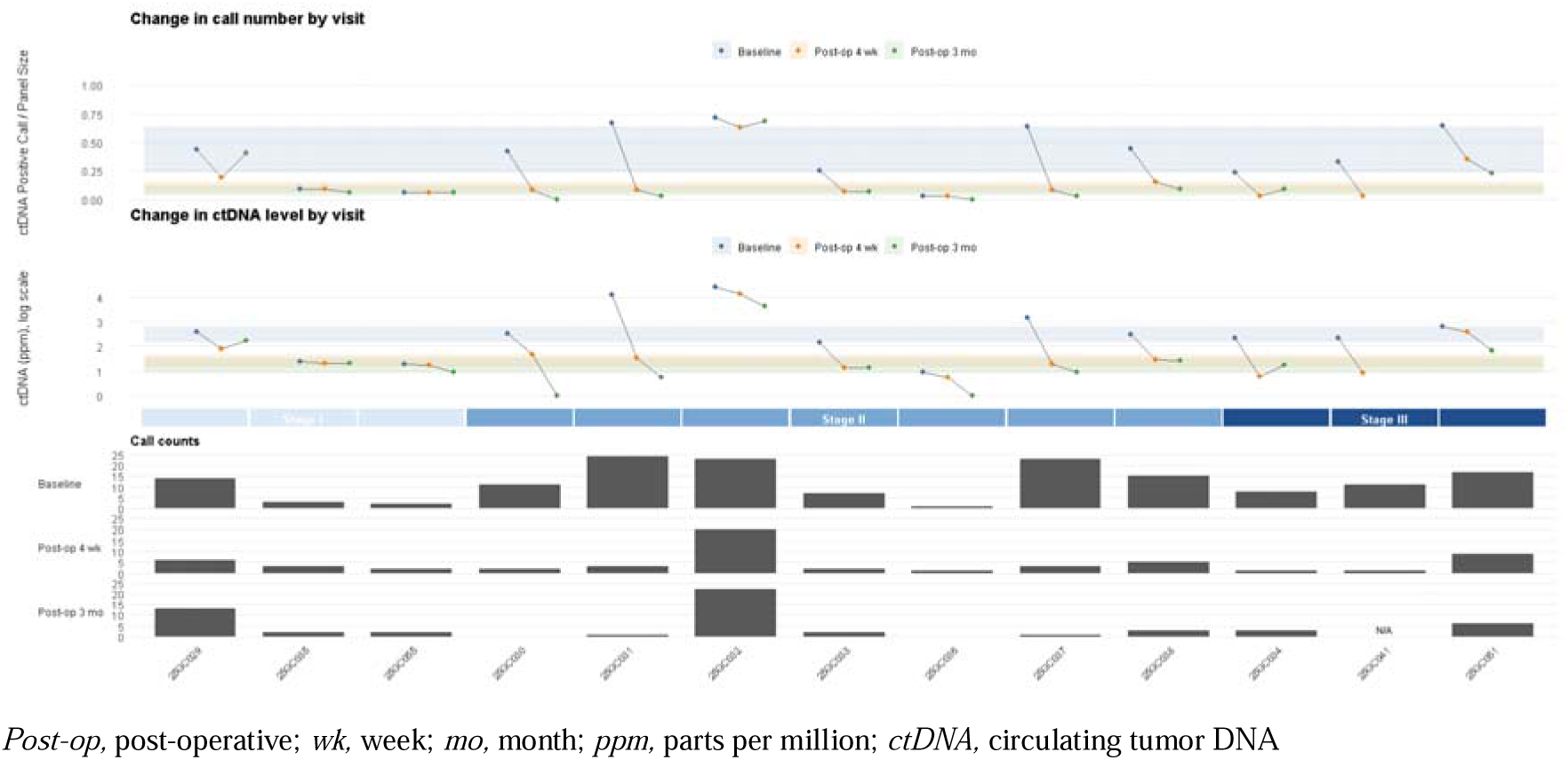
Mutation Positivity and ctDNA Level Trend. Top panels show per-patient changes in call proportion (mutation-positive call number normalized to panel size) and ctDNA particle fraction (ppm, log scale) at baseline, *Visit 1*, and *Visit 2*, ordered from left to right by pathologic stage. Bottom panels display absolute mutation-positive call counts at each time point.

Among the 10 patients with stage II–III disease, all had at least one mutation-positive call detected at *Visit 1*. Of the 9 patients with available longitudinal follow-up, 2 (22.2%) converted to no detectable MRD at *Visit 2*; both patients had two or fewer mutation-positive calls at *Visit 1*. This rate of molecular clearance is consistent with findings from studies using the Signatera™ ctDNA assay, in which 3 of 13 patients (23%) with stage III colorectal cancer achieved clearance of plasma ctDNA (*15*).

Two patients developed radiologic recurrence during follow-up. One patient with stage I disease (25GC029) had pT2N0M0 sigmoid colon adenocarcinoma with 45 regional lymph nodes examined at surgery and no nodal involvement. Tissue real-time PCR demonstrated no oncogenic mutations in *KRAS, NRAS,* or *BRAF V600*, and MSH6 expression was intact. No adverse pathological features were identified. Mutation-positive calls increased from 6 to 13 from *Visit 1* to *Visit 2*, accompanied by a rise in CPF from 79.99 ppm to 166.54 ppm. Liver metastasis was detected radiologically at 6 months and confirmed by biopsy as metastatic adenocarcinoma of colorectal origin (Supplementary Fig. S1).

Another patient with stage II disease (25GC032) had pT4N0 rectal cancer and classified as clinically high risk. Adjuvant chemotherapy was recommended but declined by the patient. The patient exhibited an increase in mutation-positive calls from 20 at *Visit 1* to 22 at *Visit 2*, and ctDNA levels remained persistently elevated (CPF 27,215.78 ppm at baseline, 14,434.80 ppm at *Visit 1*, and 4,296.01 ppm at *Visit 2*). The patient later developed radiologically confirmed local recurrence in the posterior vaginal wall 4 months after surgery. Overall, both patients who developed recurrence were MRD-positive at *Visit 1* and showed increasing mutation burden at *Visit 2,* corresponding to a median radiologic lead time of 4 months.

### Detection Behavior in Diluted Tumor DNA and Plasma cfDNA

To evaluate analytical agreement between diluted tumor genomic DNA (gDNA) and plasma cfDNA, mutation calls obtained from tumor gDNA diluted to a median of approximately 1% were compared with those detected in paired plasma samples. Overall concordance remained high across all sampling time points (Table 3), with a median of 80.5% (IQR, 73.5–93.0) at baseline, 93.0% (IQR, 60.0–100.0) at *Visit 1,* and 96.5% (IQR, 50.3–100.0) at *Visit 2*. Pairwise comparisons showed no significant differences in concordance across visits (Baseline vs. *Visit 1*, p = 0.799; Baseline vs. *Visit 2*, p = 0.838; *Visit 1* vs. *Visit 2*, p = 1.000). When stratified by pathological stage at baseline, concordance appeared numerically lower in stage I patients (median 64.5%; IQR, 50.0–84.2) than in stage II (median 89.0%; IQR, 79.0–96.5) or stage III patients (median 75.0%; IQR, 74.0–84.0), although these differences were not significant (p = 0.425), indicating stable performance irrespective of blood collection timing or disease stage.

**Table 3.** Mutational Profile Concordance Between Diluted Tumor gDNA and Plasma cfDNA.

| Pathologic Stage | Patient ID | Baseline | Visit 1 | Visit 2 |
| --- | --- | --- | --- | --- |
| **I** | **25GC029** | 79% | 83% | 92% |
|  | **25GC035** | 100% | 100% | 100% |
|  | **25GC055** | 50% | 100% | 100% |
| **II** | **25GC030** | 82% | 100% | 0%<br>(No mutation detected in cfDNA) |
|  | **25GC031** | 100% | 50% | 0%<br>(1 mutation detected in cfDNA) |
|  | **25GC032** | 93% | 93% | 93% |
|  | **25GC033** | 57% | 50% | 100% |
|  | **25GC036** | 100% | 100% | 0%<br>(No mutation detected in cfDNA) |
|  | **25GC037** | 76% | 100% | 100% |
|  | **25GC038** | 89% | 60% | 67% |
| **III** | **25GC034** | 75% | 0%<br>(1 mutation detected in cfDNA) | 100% |
|  | **25GC041** | 73% | 100% | N/A |
|  | **25GC051** | 93% | 89% | 100% |

N/A, not applicable; cfDNA, cell-free DNA

A patient with stage II disease (25GC031) had the lowest tumor burden in the cohort (30%), whereas most other patients had tumor burdens over 40%. Consistent with the relatively low tissue quality, only two variants were detected in the corresponding 1% diluted tumor gDNA sample. In contrast, plasma cfDNA analysis detected both variants and identified a total of 24 mutation-positive calls, representing 67% of the panel variants. These findings suggest that when tumor cellularity in tissue is reduced, plasma cfDNA analysis may more reliably capture tumor-derived mutations, supporting a complementary role for cfDNA when tissue quality is suboptimal. The relationship between tumor tissue quality metrics and tissue mutation-positive call rate is illustrated in Supplementary Figure S2.

### Cancer-associated Mutational Landscape Across Disease Stage

We examined the ctDNA mutational landscape of commonly reported CRC-associated genes, including *TP53, NRAS, KRAS, BRAF, APC* and *PIK3CA* (Fig. 2). In stage I disease, *APC* mutation was detected in one patient’s plasma cfDNA. In stage II, additional driver mutations were observed, including *PIK3CA, KRAS*, and *TP53*. In stage III, all three patients harbored at least one detectable cancer-associated mutation, with *TP53* mutation detected in two of three patients. This pattern parallels the canonical sequence of colorectal tumorigenesis, in which *APC* mutations typically arise early, followed by *KRAS* and *TP53* mutations as normal epithelium progresses to invasive carcinoma (*22*).

**Figure 2.**
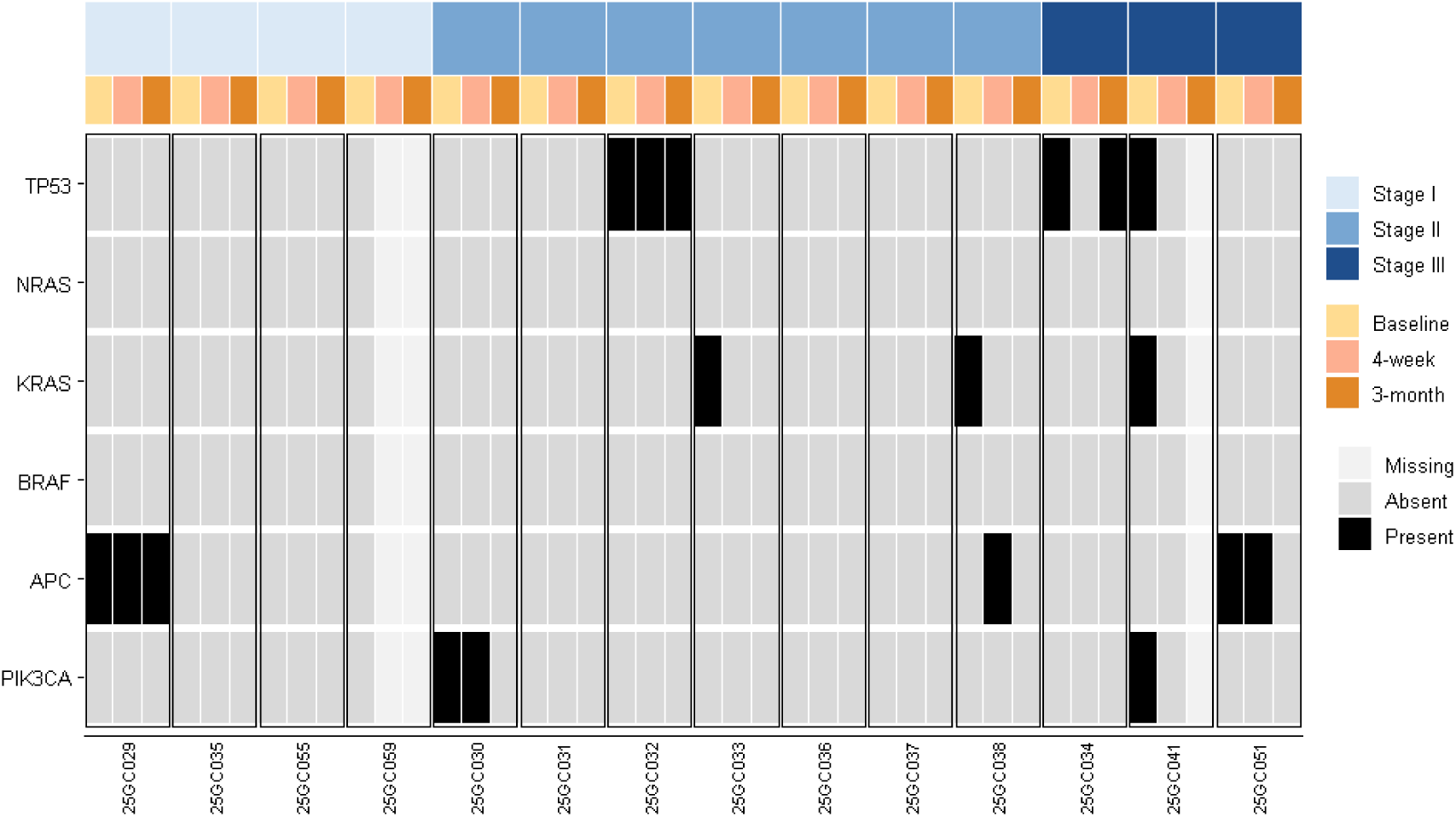
Longitudinal Detection of Key Mutations in cfDNA. Heatmap showing the presence or absence of canonical CRC-associated mutations (TP53, NRAS, KRAS, BRAF, APC, and PIK3CA) across baseline, *Visit 1,* and *Visit 2*, stratified by pathological stage. Mutation detection demonstrates a stage-dependent pattern at baseline with overall reduction in post-operative detectability.

Following surgical resection, these CRC-associated mutations showed a marked decline in detectability. When the maximum VAF among the six canonical driver genes was evaluated per patient, a significant postoperative decrease was observed (Supplementary Fig. S3). Median maximum VAF declined from 0.052% (IQR, 0.000–0.084) at baseline to 0.000% (IQR, 0.000–0.025) at *Visit 1* (p = 0.021). Although a further reduction was observed between *Visit 1* and *Visit 2*, this change did not reach statistical significance (p = 0.208), suggesting that the major decrease in driver mutation burden occurred early after surgical resection. Two patients demonstrated an increase in maximum VAF of key CRC-associated mutations at *Visit 2* compared with *Visit 1* (25GC029, stage I; 25GC034, stage III), accompanied by increases in mutation-positive call number and CPF, indicating concordant upward trends across molecular metrics. One of these patients (25GC029) developed recurrence within 6 months after surgery, whereas the other remains recurrence-free to date. To sum it up, canonical CRC driver mutations were detectable in a stage-dependent manner pre-operatively and their VAFs declined substantially after surgery, although a subset of patients exhibited early quantitative rebound requiring close surveillance.

### Impact of Analytical Sensitivity on Post-operative MRD Positivity

Conventional ctDNA MRD assays typically report limits of detection around 0.01% VAF (approximately 100 ppm), with MRD detection primarily occurring above this range (*23*). Since post-operative ctDNA levels spanned a wide dynamic range across individuals, we evaluated how different hypothetical analytical detection thresholds would affect the observed MRD positivity rate in the stage II–III cohort. In our cohort, the lower bound of detectable ctDNA reached 8.26 ppm at baseline, 4.67 ppm at *Visit 1*, and 4.92 ppm at *Visit 2*. Using progressively lower thresholds (100, 40, 30, 20, 10, and 1-ppm), post-operative MRD positivity was calculated at both follow-up visits (Fig. 3). At *Visit 1*, 20% of patients were classified as MRD-positive at a threshold of 100 ppm. This proportion increased to 30% at 40 ppm, 40% at 30 ppm, 50% at 20 ppm, 70% at 10 ppm, and 100% at 1 ppm. In the NCCTG N0147 (Alliance) trial analyzing stage III CRC using Guardant Reveal™ (Guardant Health), 20.4% of patients were ctDNA-positive after surgery and before adjuvant chemotherapy (*24*); our cohort demonstrated a similar magnitude of detection under comparable analytical sensitivity assumptions. Among the 10 patients evaluated at *Visit 1*, 8 patients (80%) harbored detectable ctDNA levels below 100 ppm. Of these patients, four (50%) had levels below 15 ppm. This distribution aligns with recent findings from the NeXT Personal® Dx (NPDx) ctDNA MRD assay, which reports a detection threshold of approximately 1.67 ppm and observed that 50.4% of ultrasensitive detections across multiple cancer types occurred at or below 15 ppm (*25, 26*).

**Figure 3.**
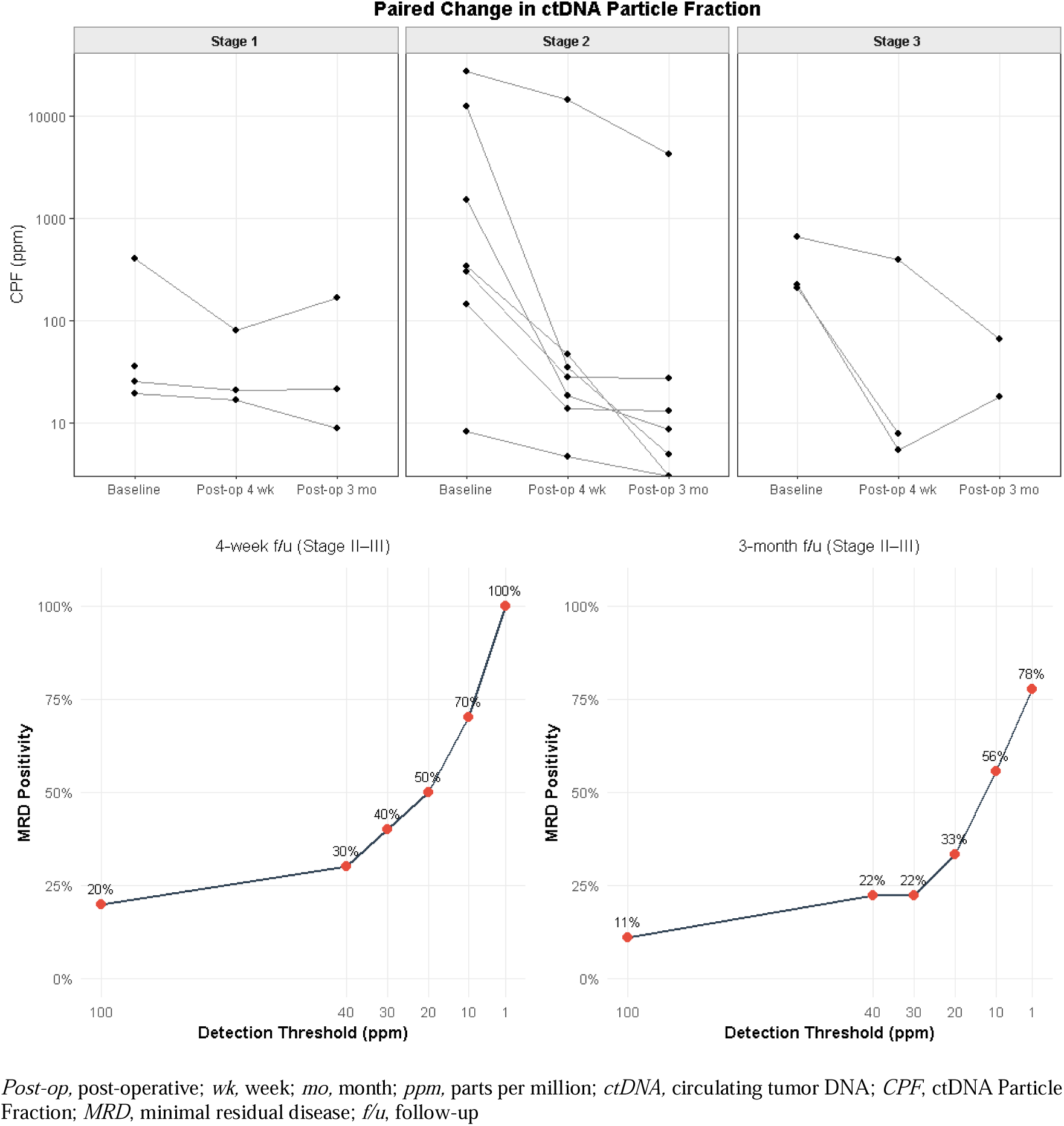
Threshold-dependent MRD Positivity After Curative Surgery. Top panels show paired changes in ctDNA particle fraction (CPF, ppm; log scale) across baseline, *Visit 1* (post-op 4-week), and *Visit 2* (post-op 3-month) by pathologic stage. Bottom panels show MRD positivity rates in stage II–III patients at *Visit 1* and *Visit 2* under progressively lower hypothetical detection thresholds (100 to 1 ppm).

A similar pattern was observed at follow-up. MRD positivity was 11% at the 100 ppm detection threshold, increasing to 22% at 40 ppm, 33% at 20 ppm, 56% at 10 ppm, and 78% at 1 ppm. Among the 7 stage II–III patients classified as MRD-positive at *Visit 2*, 6 patients (85.7%) had ctDNA levels below 100 ppm, falling within the ultrasensitive range. Half of these patients (n = 3) also had ctDNA levels below 15 ppm. These findings suggest that post-operative MRD classification is strongly influenced by analytical sensitivity, as a notable proportion of positive cases occurs at ctDNA levels below 15 ppm. Assays with higher detection thresholds may classify patients with residual disease as MRD-negative. Accordingly, analytical performance characteristics should be taken into consideration when interpreting MRD findings and when comparing results across platforms.

## Discussion

In this prospective pilot study of patients with stage I–III CRC undergoing curative-intent surgery, we evaluated the performance of a CRISPR-enhanced plasma sequencing assay, MUTE-Seq, for MRD detection. All patients had detectable ctDNA at baseline, and the mutation-positive call number declined most markedly within 4-week following surgical resection. Canonical CRC driver mutations were detected in a stage-dependent manner at baseline and exhibited substantial quantitative reductions in VAF post-operatively. Analytical concordance between diluted tumor genomic DNA and plasma cfDNA remained consistently high, increasing from a median of 80.5% at baseline to 93.0% at *Visit 1* and 96.5% at *Visit 2.* High concordance at low variant allele fractions supports the analytical reliability of the CRISPR-based approach in the MRD setting.

Most notably, MRD positivity rates were highly dependent on analytical sensitivity. In the Alliance trial, approximately 20% of patients were ctDNA-positive after surgery at a reported detection threshold around 0.01% VAF (*24*). When applying a comparable 100 ppm threshold in our dataset, MRD positivity at *Visit 1* was similarly 20%, demonstrating concordance with published rates under equivalent sensitivity assumptions. However, when lower thresholds were applied, MRD positivity increased substantially. At *Visit 1,* 80% of MRD-positive patients with stage II–III disease harbored ctDNA levels below 100 ppm, and half of these cases were below 15 ppm. A similar distribution was observed at *Visit 2,* where 85.7% of MRD-positive stage II–III patients had ctDNA levels below 100 ppm, and half again fell below 15 ppm. Reports from ultrasensitive platforms, including NeXT Personal® Dx, have shown that a sizable proportion of ctDNA-positive cases occur below 15 ppm (*25*). These findings indicate that post-operative MRD prevalence is a biological parameter strongly shaped by the detection threshold of the assay, and that residual disease signals frequently exist at levels lower than the conventional 0.01% VAF limit. Interpretation of MRD-negative results must therefore account for analytical limits of detection, as insufficient sensitivity may underestimate true residual disease burden.

Now the clinical question extends beyond binary MRD positivity or negativity. When a hypothetical detection threshold of 1 ppm was applied, MRD positivity at the 3-month landmark in stage II–III CRC reached 78% in this study; it is unlikely that all of these patients will recur. According to the ACCENT database, the recurrence rate for stage II–III colon cancer is approximately 32.9% (*27*). The field of liquid biopsy may be entering an era of quantitative MRD interpretation. An instructive parallel exists in prostate cancer, where biochemical recurrence is defined using prostate specific antigen (PSA) level rather than mere detectability. According to American Urological Association criteria, biochemical recurrence after prostatectomy is defined as a PSA ≥0.2 ng/mL confirmed by a second measurement, whereas American Society for Therapeutic Radiology and Oncology defines it as a rise of ≥2.0 ng/mL above nadir after radiotherapy (*28, 29*). These thresholds reflect clinical validation linking quantitative biomarker levels and trajectories to recurrence risk. It is therefore conceivable that future definitions of molecular recurrence will integrate longitudinal trend and quantitative mutation burden rather than relying solely on presence or absence of signal. Establishing cutoffs may represent the next critical step in MRD-guided management.

The sample size of this study is small and findings are preliminary. Follow-up duration remains limited, precluding definitive conclusions regarding long-term prognostic performance. While our data suggest that quantitative ctDNA levels and mutation-positive call numbers may approximate true recurrence risk, larger prospective studies are warranted to define clinically meaningful ctDNA cutoffs and to determine how ultrasensitive detection can be translated into improved risk stratification and therapeutic guidance in the adjuvant setting.

## Materials and Methods

### Data Ethics Statement

This prospective study was conducted in accordance with the principles of the Declaration of Helsinki and was approved by the Institutional Review Board of Korea University Anam Hospital (IRB No. 2022AN0090). All participants received a detailed explanation of the study objectives and procedures and provided written informed consent prior to enrollment. Patients were enrolled from June to July 2025, with clinical data and biospecimens collected through December 2025 at Korea University Anam Hospital.

Eligible participants were adults aged 19 years or older with a histologically confirmed diagnosis of cancer or with suspected malignancy undergoing diagnostic evaluation, surgical intervention, or cancer-related testing or treatment. Patients were excluded if they were pregnant, younger than 19 years, or unable or unwilling to provide written informed consent. Additional exclusion criteria included a diagnosis of any hematologic malignancy or solid tumor within the past 5 years, rare diseases or hereditary disorders, a diagnosis of inflammatory bowel disease including Crohn’s disease or ulcerative colitis, or suspected hereditary colorectal cancer syndromes such as familial adenomatous polyposis or hereditary nonpolyposis colorectal cancer. Study participation was terminated if adequate biospecimens could not be obtained, or if the participant withdrew consent.

### Specimen Collection and Molecular Analysis

Tumor tissue samples were obtained at the time of surgery and processed in the pathology department to generate formalin-fixed paraffin-embedded blocks. Sixteen slides were sectioned from each block. One slide was stained and used as a guide slide to annotate tumor regions. Based on this annotation, tumor areas from the remaining 15 slides were macrodissected and used for genomic DNA extraction prior to sequencing analyses. Peripheral blood (20 mL) was collected into cfDNA-stabilizing tubes at three predefined time points: preoperatively (baseline) and post-operatively at the first (4-week) and second (3-month) follow-up visits. None of the stage II patients received adjuvant chemotherapy, whereas all stage III patients were treated with adjuvant FOLFOX. For stage III patients, the 4-week post-operative blood sampling was performed at the initiation of adjuvant chemotherapy, and the 3-month sampling was obtained after approximately 2 months of adjuvant treatment. Blood samples were centrifuged on the day of collection to separate plasma and buffy coat, which were stored at −80 °C until cfDNA and genomic DNA extraction, respectively. Plasma ctDNA sequencing was performed using the MUTE-Seq assay (GeneCker), as previously described (*21*). The overall experimental workflow is illustrated in Figure 4. Baseline plasma and matched tumor tissue samples were available for 14 patients. Following withdrawal of consent, 13 patients were evaluable at *Visit 1* and 12 at *Visit 2*.

**Figure 4.**
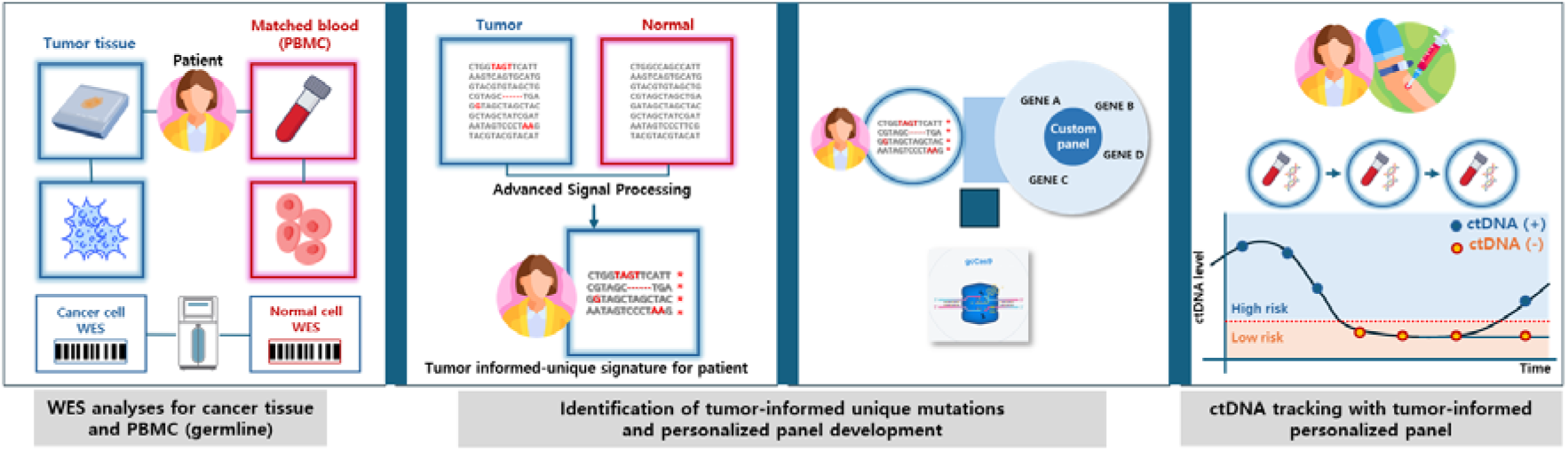
Overview of the Tumor-informed MRD Workflow. Whole exome sequencing (WES) of tumor tissue and matched peripheral blood mononuclear cells (PBMCs) is performed to identify patient-specific somatic mutations, which are incorporated into a customized panel for longitudinal monitoring of circulating tumor DNA (ctDNA) through serial blood sampling.

The median panel size was 33 loci. To summarize tumor-derived signals at the sample level, we defined the ctDNA particle fraction as the normalized sum of locus-specific VAFs across targetable variants included in the sequencing panel, expressed in ppm. For each locus, the number of sequencing reads supporting the mutant allele was divided by the total number of reads covering that genomic position to calculate the VAF. These VAF values were then summed across all loci that returned a positive mutation call and divided by the total number of panel loci for which successful amplicon formation was achieved. The resulting value was multiplied by 1,000,000 to obtain the final CPF in ppm. Samples without detectable variants were assigned a CPF of zero. This normalization adjusted for differences in panel breadth. Potential germline variants were excluded from the panel using matched normal DNA derived from peripheral blood cells.

To assess analytical agreement under controlled low–variant allele fraction conditions, tumor tissue genomic DNA was diluted with matched PBMC genomic DNA to achieve a median VAF of approximately 1%. Mutation calls obtained from these diluted samples were compared with those detected in paired plasma cfDNA analyzed using the same sequencing panel. Concordance was defined as the number of shared mutation-positive calls between the diluted tissue sample and plasma cfDNA, normalized to the smaller number of positive calls observed in either sample.

### Statistical Analysis

Continuous variables were summarized using the median as a measure of central tendency, with variability described by the IQR. Group comparisons were performed using the Wilcoxon rank-sum test, the *t* test or the Kruskal-Wallis test, depending on data distribution and the number of groups compared. Categorical variables were analyzed using the χ² test or Fisher’s exact test. All statistical analyses were conducted using R version 4.5.1 (R Project for Statistical Computing). A two-sided *p* value < 0.05 was considered statistically significant. Figures were generated using the ggplot2 package (*30*).

## Data Availability Statement

The data generated in this study are available upon reasonable request from the corresponding author.

## Notes

### Competing Interest Statement

Hong-Kyu Kim and Jin-Soo Kim report grants and personal fees from GeneCker during the conduct of the study. Patents describing technologies used in this study have been obtained by Sunghyeok Ye.

### Funding Statement

This study was funded by Korea University Medicine and Seoul Clinical Laboratories (SCL) Grant.

### Author Declarations

Institutional Review Board of Korea University Anam Hospital gave ethical approval for this work (IRB No. 2022AN0090).

