## Supplementary Fig. S1 for "Beyond Binary MRD: Quantitative ctDNA Interpretation After Curative-Intent Surgery for Colorectal Cancer"

**Supplementary Figure S1. Abdominal Imaging of Patient 25GC029 with Stage I Colon Cancer and Subsequent Recurrence**

(Left) Pre-operative evaluation with PET/CT and contrast-enhanced abdominal CT showing no evidence of distant metastasis. (Right) PET/CT and abdominal MRI at 6 months after surgery demonstrating a newly developed hepatic lesion with a size of 9mm consistent with metastatic recurrence.

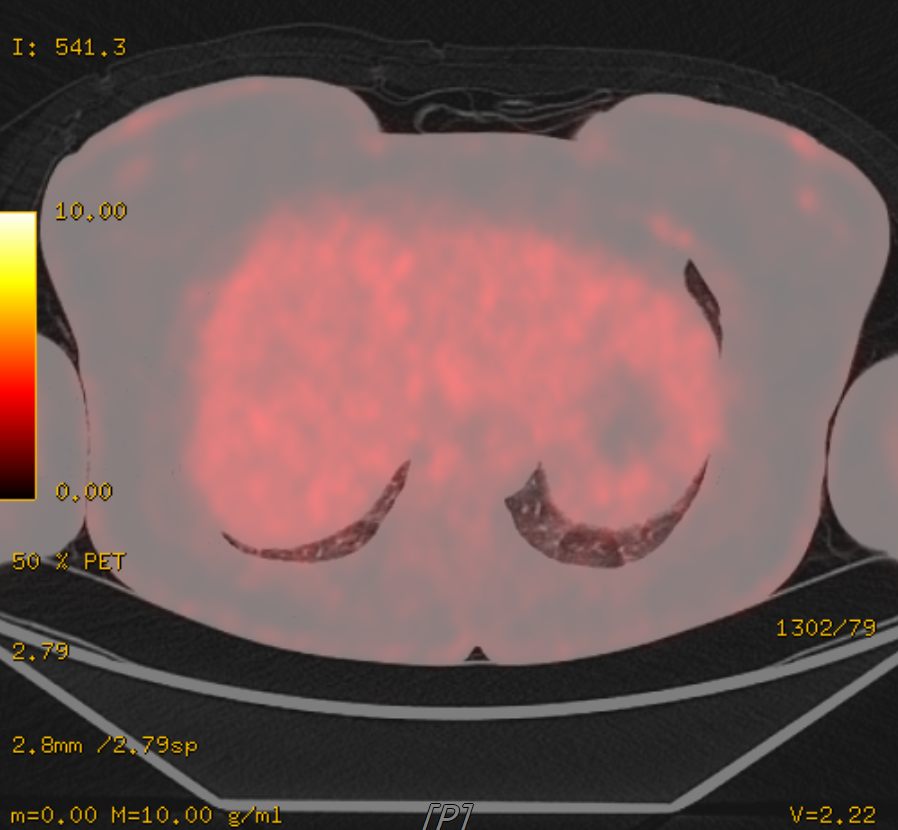

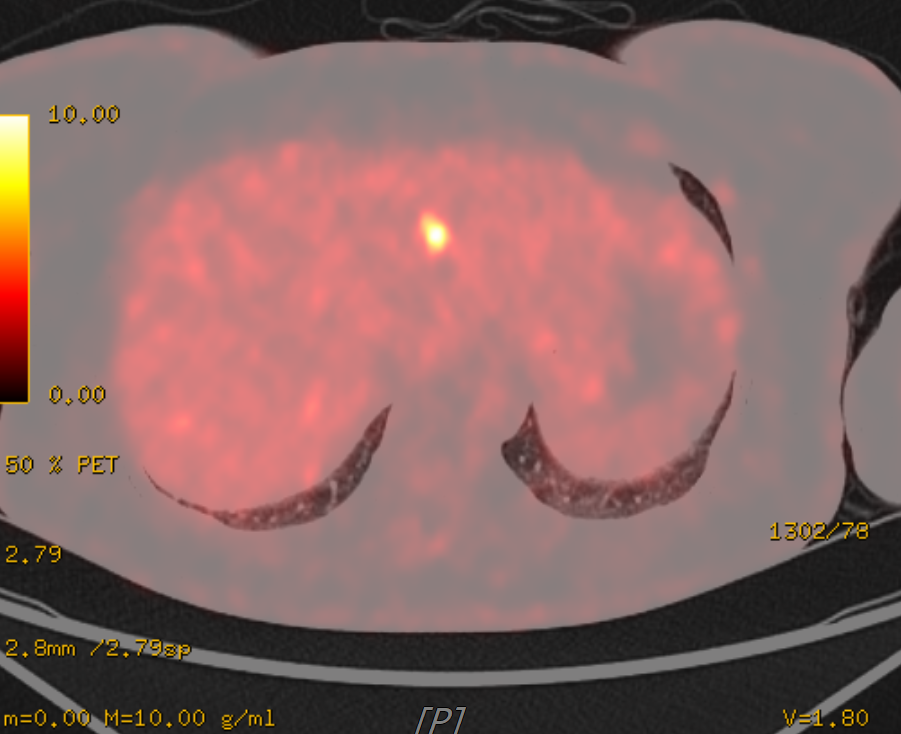

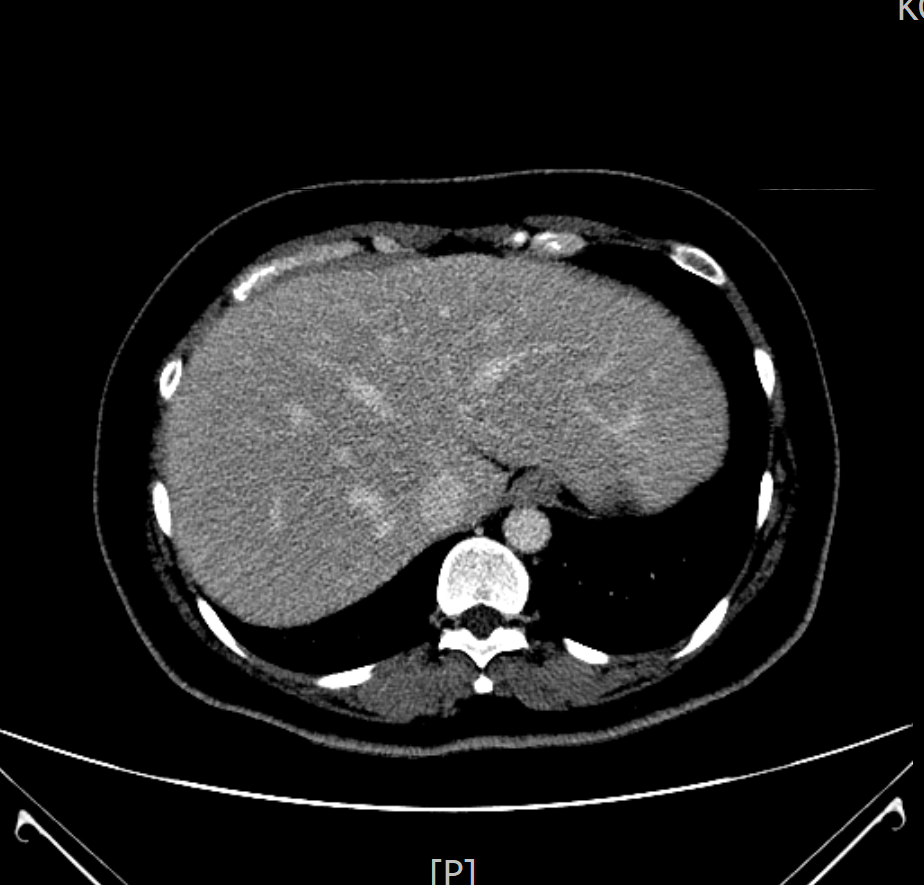

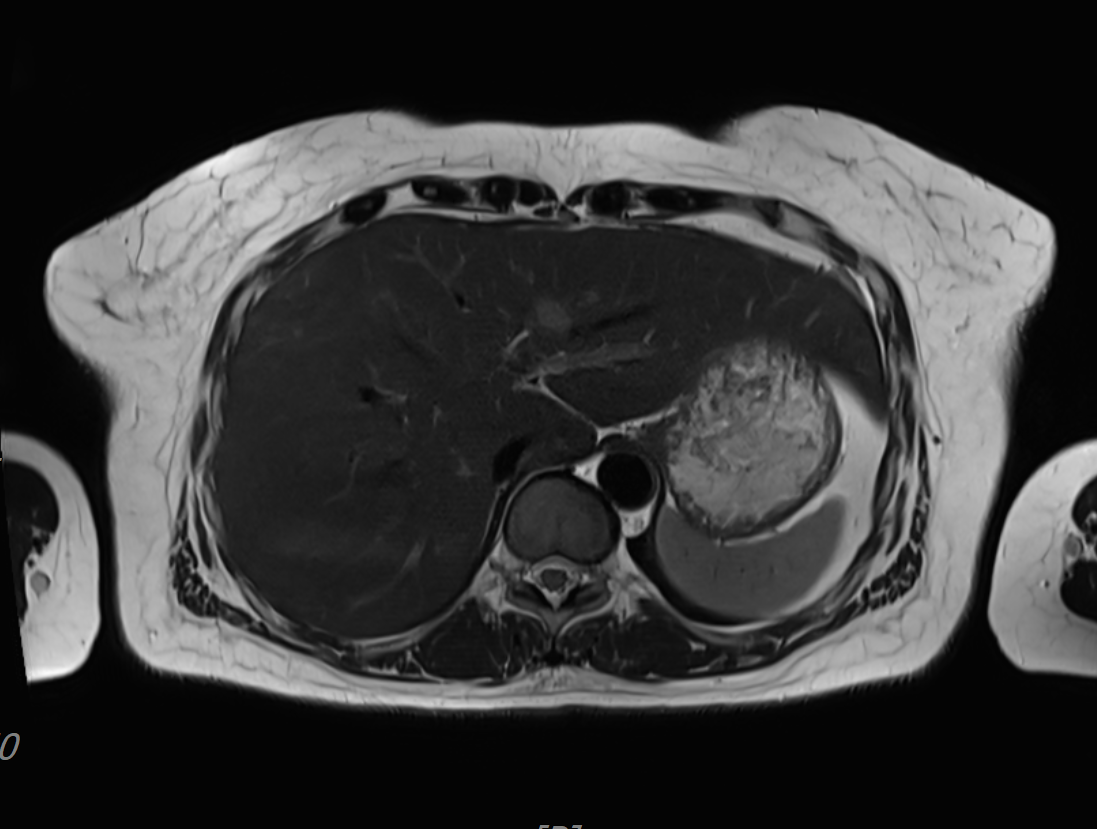
