## Supplementary Figure S2 for "Beyond Binary MRD: Quantitative ctDNA Interpretation After Curative-Intent Surgery for Colorectal Cancer"

**Supplementary Figure S2. Tumor Tissue Mutation Detection Rate and Tissue Quality Metrics**

Scatter plots showing the relationship between tumor tissue mutation-positive call proportion and **(left)** tumor cellularity (%) and **(right)** tumor genomic DNA integrity number (DIN). No significant correlation was observed; patient 25GC031 exhibited a low tissue mutation detection rate and had the lowest tumor burden among the cohort.

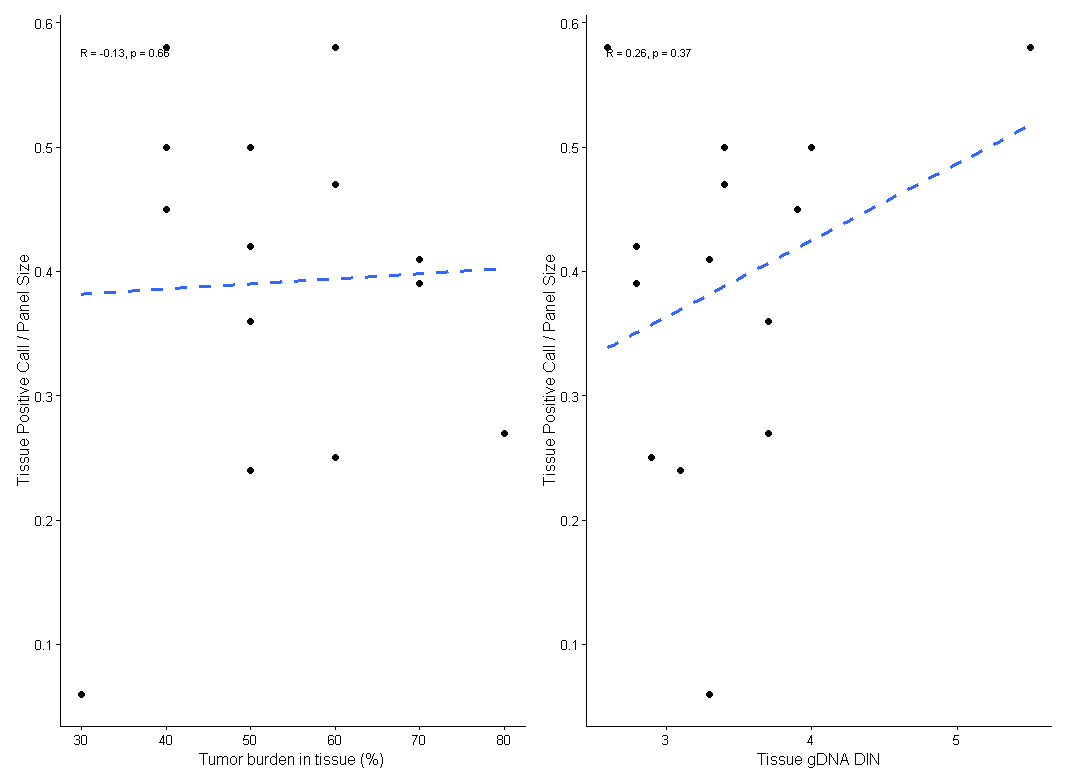

25GC031

*gDNA*, genomic DNA; *DIN*, DNA integrity number
