## Supplementary Fig. S3 for "Beyond Binary MRD: Quantitative ctDNA Interpretation After Curative-Intent Surgery for Colorectal Cancer"

**Supplementary Figure S3. Postoperative Reduction in Maximum Key Mutation VAF**

Per-patient changes in the maximum variant allele fraction (VAF) among six key CRC-associated gene mutations (*TP53, NRAS, KRAS, BRAF, APC*, and *PIK3CA*) across baseline, *Visit 1* and *Visit 2* (log scale).

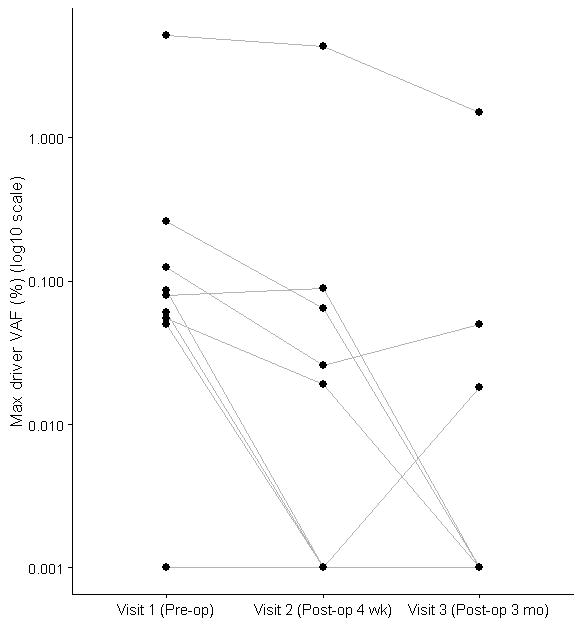

Baseline	Visit 1	Visit 2

*Max,* maximum; *VAF,* variant allele fraction; *pre-op,* pre-operative; *post-op,* post-operative; *wk,* week; *mo,* month.
